# “Does a respiratory virus have an ecological niche, and if so, can it be mapped?” Yes and yes

**DOI:** 10.1101/2022.05.04.22274675

**Authors:** Christopher R. Stephens, Constantino González-Salazar, Pedro Romero Martínez

## Abstract

Although the utility of Ecological Niche models (ENM) and Species Distribution models (SDM) has been demonstrated in many ecological applications, their suitability for modelling epidemics or pandemics, such as SARS-Cov-2, has been questioned. In this paper, contrary to this viewpoint, we show that ENMs and SDMs can be created that can describe the evolution of pandemics, both in space and time. As an illustrative use case, we create models for predicting confirmed cases of COVID-19, viewed as our target “species”, in Mexico through 2020 and 2021, showing that the models are predictive in both space and time. In order to achieve this, we extend a recently developed Bayesian framework for niche modelling, to include: i) dynamic, non-equilibrium “species” distributions; ii) a wider set of habitat variables, including behavioural, socio-economic and socio-demographic variables, as well as standard climatic variables; iii) distinct models and associated niches for different species characteristics, showing how the niche, as deduced through presence-absence data, can differ from that deduced from abundance data. We show that the niche associated with those places with the highest abundance of cases has been highly conserved throughout the pandemic, while the inferred niche associated with presence of cases has been changing. Finally, we show how causal chains can be inferred and confounding identified by showing that behavioural and social factors are much more predictive than climate and that, further, the latter is confounded by the former.

## 1 Introduction

Recently there has been debate [1, 2, 3, 4, 5, 6] as to whether Species Distribution Models (SDM) are appropriate tools in the study of the COVID-19 pandemic. Of course, such a debate raises the important question of when and under what circumstances an SDM, or an Ecological Niche model (ENM), are likely to be valid and/or useful in the study of disease in general? Just which diseases, or aspects of diseases, can be usefully studied using SDM/ENMs? Which pathogens have ecological niches and which do not? And, if some don’t, why don’t they? Taking the broad view: If ecology is the study of the relations between organisms, both among themselves and with their environment, and an ecological niche is the full set of biotic and abiotic factors that favour the presence of an organism [7, 8], then clearly all disease pathogens must have an ecological niche. The question, rather, is: how is that niche to be characterised and quantified? What are the appropriate niche dimensions and can a meaningful and useful ENM and, consequently an SDM, be constructed?

The application of ENM/SDMs to disease has been principally to zoonoses [9, 10], where non-human species are involved in the transmission cycle. In this case, both abiotic and biotic factors have been considered as niche variables [11] and different agents of the transmission cycle itself, including the pathogen, its vectors and its hosts as target variable [12]. Due to methodological shortcomings, however, the large majority of these studies have used only abiotic niche factors, with models constructed with the implicit assumption that the system was in “equilibrium” [13].

A further question is: when is an ecological versus an epidemiological approach more appropriate? The difference between the two is stated quite clearly in the epidemiological literature [14, 15]. Put simply, ecological models are naturally associated with the “where” of a disease, while epidemiological models are more concerned with the “who”. This difference is aptly captured in [16] with the question: “Is it better to have a heart attack in the United States or Canada?”, the emphasis being that this type of question is of an ecological nature, whereas the question: How many people out of a cohort of individuals diagnosed with COVID-19 will subsequently die? is an epidemiological question. Indeed, this ecological “where” perspective has a long history in the social sciences as a whole [17, 18]. From a modelling viewpoint, the difference between the two approaches lies in what statistical ensemble will be used to draw inferences. Epidemiological models use an ensemble of “individuals”, while ecological models use an ensemble of “places”, where each place is associated, either explicitly or implicitly, with a population. The “places” can vary depending on context: from countries and other political units, to eco-regions, any fixed-area spatial cells, and to pixels on a raster.

Additionally, there is a subsequent question of “when”, that is relevant for both the ecological and epidemiological points of view. Standard SIR-type modelling [19], for instance, is associated with the “who” and the “when”, with the “who” being associated with a fixed-number of states -susceptible, infected etc. that are associated with individuals. These models can be extended to consider “place” by constructing a SIR-type model for a particular place [20]. Such models, however, do not account for the distinguishing features of that particular “place”, as does an SDM/ENM. On the other hand, currently, SDM/ENMs do not account for the dynamics of a disease and are also unnecessarily restrictive in the set of niche variables considered. Finally, there is, behind the “who”, “where” and “when”, the “why” that explains them. This we argue is the most important role for the use of ENMs in the study of human disease.

In this paper we show that an ecological approach, using SDM/ENMs, can be usefully applied to any human or non-human disease, transmissible or not, taking as a specific example the spatio-temporal distribution of COVID-19 in Mexico to answer in the affirmative that a respiratory virus does, indeed, have an ecological niche, and that it can be mapped. We show that a wide variety of habitat variables, both environmental, behavioural and social, and of different types and spatio-temporal resolutions, can be included, to yield a deeper understanding of the factors that drive the COVID-19 pandemic. Moreover, we show how to generalise SDM/ENMs to incorporate and predict the dynamics of the disease. Finally, we show how the formalism can be used to disentangle the complex causal chains that are a fundamental part of a Complex Adaptive System.

## 2 Results

An important aspect of a complex, adaptive phenomenon, such as the COVID-19 pandemic, is that there are many questions of “where”, “when” and “why” that require answers, with each question in turn requiring its own SDM/ENM. Here, we will present representative results for the following: i) Where are the highest number of confirmed cases in a given time period to be found? ii) Where are confirmed cases in a given time period to be found? iii) Where will most confirmed cases be found in a future time period? iv) Where will cases be found in a future time period where previously there were none? In a more ecological language and taking confirmed cases of COVID-19 as the “species” of interest: i) Where is the highest abundance of the species of interest to be found? ii) Where is the species to be found in a given time period independently of its abundance? iii) Where will the species be found in highest abundance in a future time period? iv) Where will the species be found where it was not present previously? ^1^ These five representative models illustrate important differences about how ENM/SDMs can be used to answer where, when and why questions. Firstly, we may use a model developed on a region *R*_*train*_(*t*), using data from a time period *t*, to predict on another region *R*_*test*_(*t*), as is standardly done, and where the complete spatial region under consideration is *R*(*t*) = *R*_*train*_(*t*)∪*R*_*test*_(*t*). Secondly, we may use a model developed on a region *R*(*t*) to predict on *R*(*t*′), the same region at some later time.

### 2.1 Dynamic biotic and abiotic factors can be included in an SDM/ENM for COVID-19

To answer each of the above questions, we must extend standard ENM/SDM modelling to include both dynamical target classes, *C*(*t*), and associated dynamical habitat variables, **X**(*t*′), and develop SDM/ENMs, *P* (*C*(*t*) | **X**(*t*′)), that relate them. With question i) the target class will be the 10% of Mexican municipalities that have the highest number of cases in month *t*. In that context we are characterising niche by the relative abundance of the target species, with those locations where it is highest being characterised as most niche-like. This target class is time dependent, as it may be that those places with the highest abundance at one time are not the same as those at another. With question ii) the target class will be presence of confirmed cases in month *t*. Questions iii) and iv) refer to these target classes but in a future month *t*′. In particular, for presence of confirmed cases we will be interested in predicting those municipalities that had no previous cases in time period *t* but do in period *t*′.

Thus, the targets for our ENM/SDMs are all, in principle, time-dependent. For the habitat variables, we use those presented in section 4.7 below, with an illustrative dynamic habitat variable being the decile of municipalities corresponding to a given range of number of confirmed cases in the period *t* − 1.

### 2.2 Predictive SDM/ENMs can be created for COVID-19

SDM/ENMs were created for the target classes i)-v). In Figure 1 (left) we see the results for a purely spatial ENM/SDM model with target class, corresponding to question i), being the 10% of municipalities with the highest number of confirmed cases, considered for three different, represnetative months of the pandemic: March 2020, June 2020 and January 2021. In this case, performance is based on a 5-fold 70%/30% train/test split of the 2458 municipalities. Figure 1 (left) shows the corresponding ROC curves and the corresponding AUC for five separate SDM/ENMs for each test set, with: socio-demographic/economic variables only, mobility only, climate only, air contamination only and all factors together, for each of the three considered months. The relative performance of each sub-model is clear, with the mobility and socio-demographic models being the best performers, followed by the air contamination model, then the climate model, with the Total model equivalent to the mobility and socio-demographic models. Table 2 shows the most niche-like and most anti-niche-like habitat factors, as ranked by the statistical measure of co-occurrence *ε* (see the Methods section) for each month. As can be seen, the top 18 and bottom 18 ranked factors are almost identical for the three different periods. In

**Figure 1:**
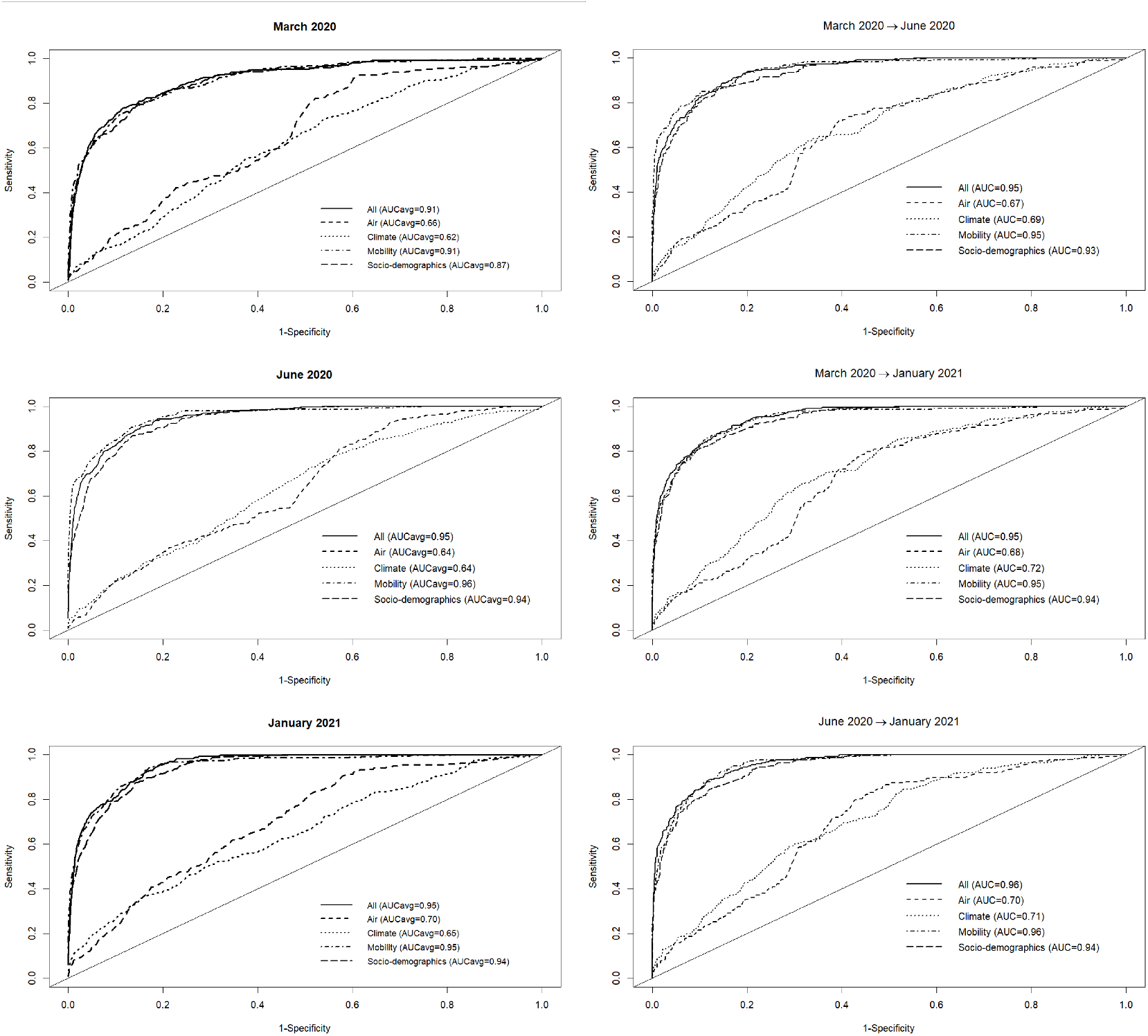
Left: Performance for 5 different SDMs for the class - top 10% of municipalities with the highest number of cases -according to the habitat variable group considered (mobility, socio-demographics, air, climate and all) for three different months of the pandemic. Five different train-test 70%-30% splits were considered for each month; Right: Performance for 5 different SDMs for the class - top 10% of municipalities with the highest number of cases - according to the habitat variable group considered (mobility, socio-demographics, air, climate and all) using a model trained on one month and tested on a future month.

Figure 3 (bottom) we show the values of the score 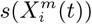 (see the Methods section) for the 10 deciles, *m* = [1, 10], of the habitat variable *X*_*i*_ =*Internal labor flow of the municipality*, observing that the score contributions are highly conserved across the three time periods. This niche conservation effect is shown in an even more striking fashion in Figure 4, where we see the correlation between the 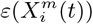 values (left) and the score values, 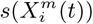, (right) for the ENM models for each individual habitat variable for the three different periods. For instance, we observe that the 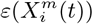 values from March 2020 explain 96% of the variance in the 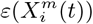 values for January 2021. As a further test of niche conservation, we used an ENM trained on all of the cells in one month to predict a future month. In Figure 1 (right), we see the ROC and AUC for the 5 models using different categories of habitat variable (mobility, socio-demographic, air contamination, climate and all) trained on data from all spatial cells for the period March 2020 and tested on the periods June 2020 and January 2021. Similarly, we show the result of analogous models trained on the period June 2020 and tested on the period January 2021.

**Figure 2:**
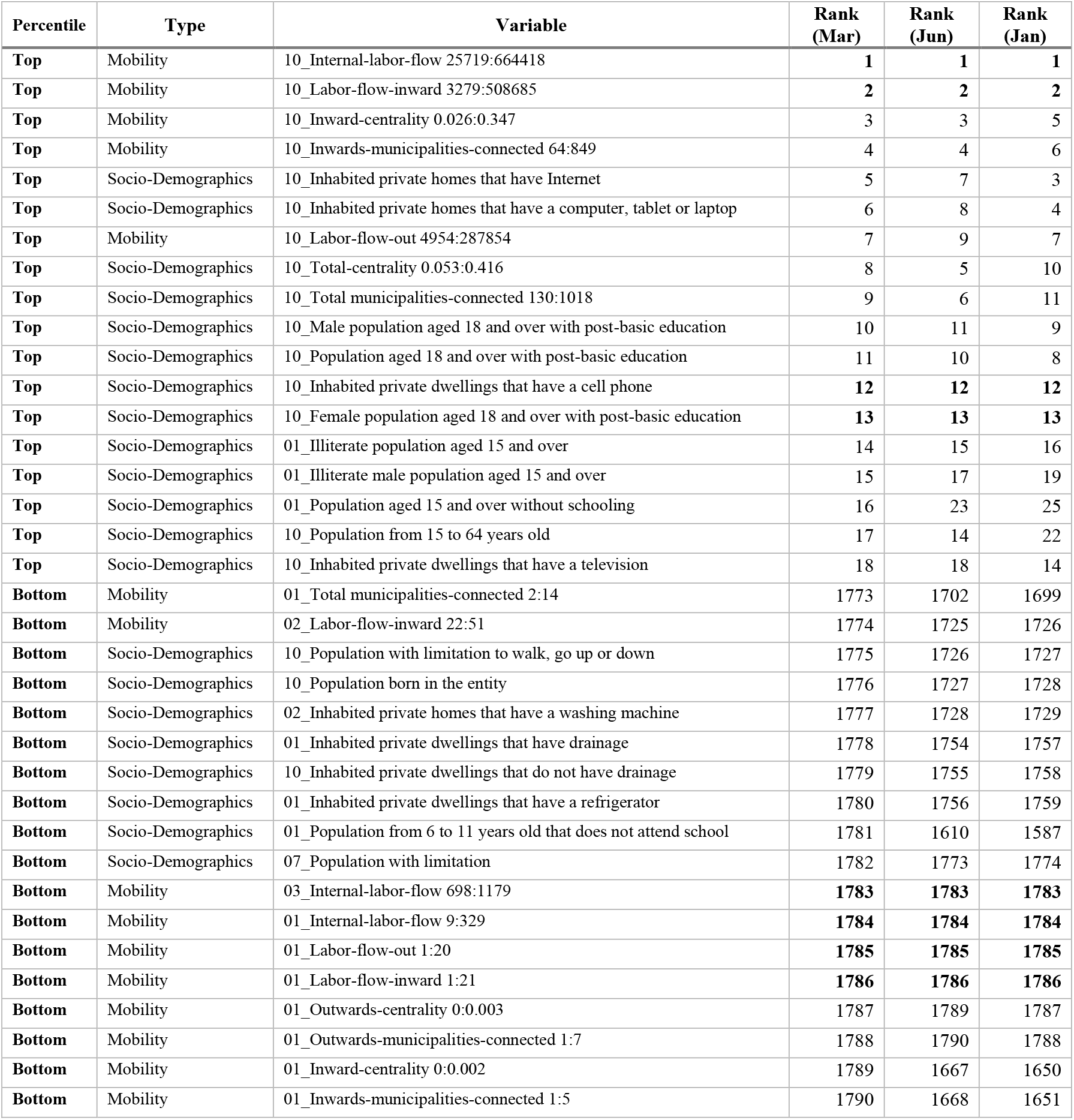
Top 18 most niche-like and bottom 18 most anti-niche-like habitat factors for the ENM for top 10% of municipalities with highest number of cases ranked by score in March 2020 and showing the corresponding rank in June 2020 and January 2021.

**Figure 3:**
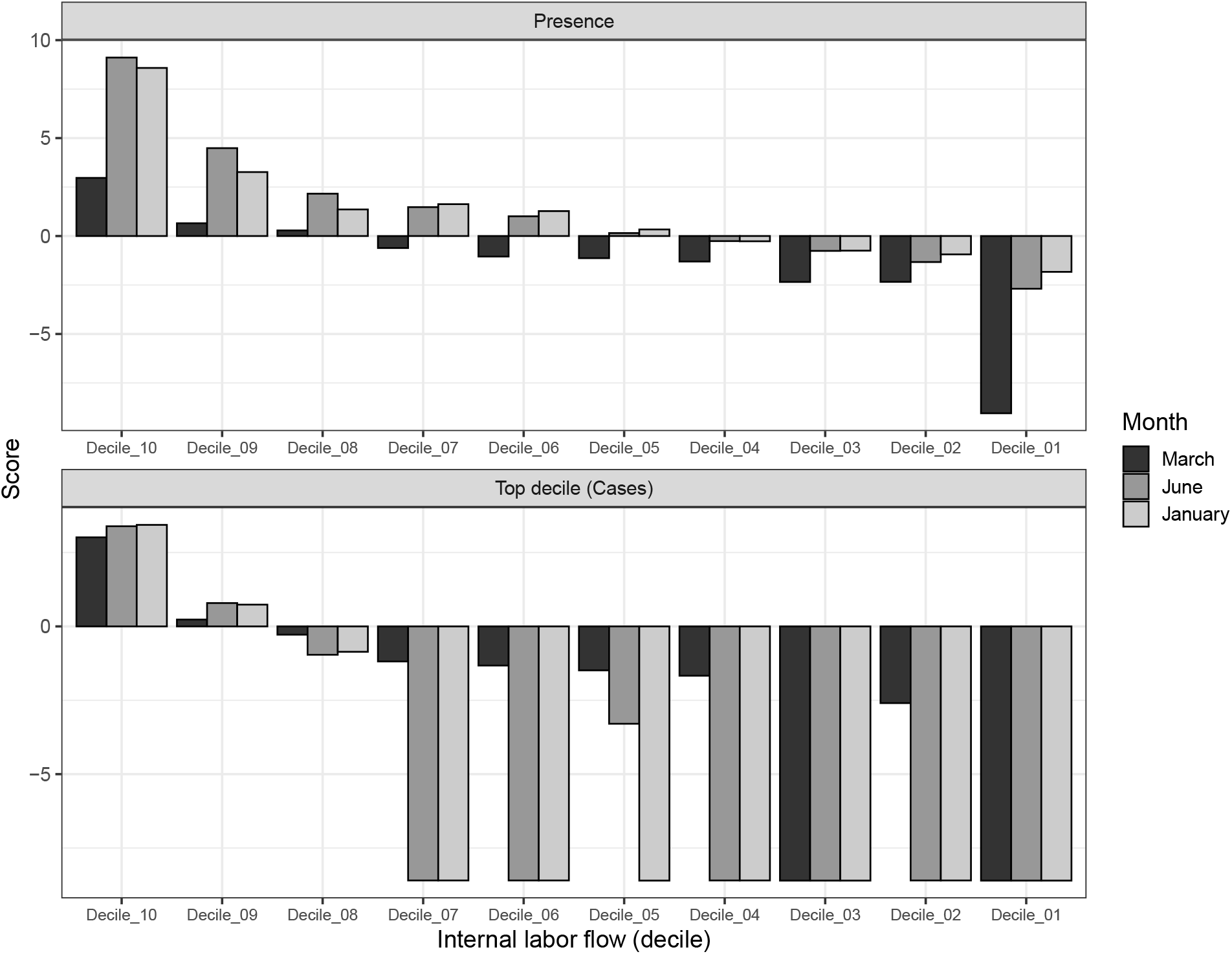
Top: Model scores for the 10 deciles (*Decile*_*i*_) of the habitat variable *Internal labor flow of the municipality* for the model predicting presence of COVID-19 cases; Bottom: Model scores for the 10 deciles (*Decile*_*i*_) of the habitat variable *Internal labor flow of the municipality* for the model predicting the top 10% of municipalities with highest number of cases.

**Figure 4:**
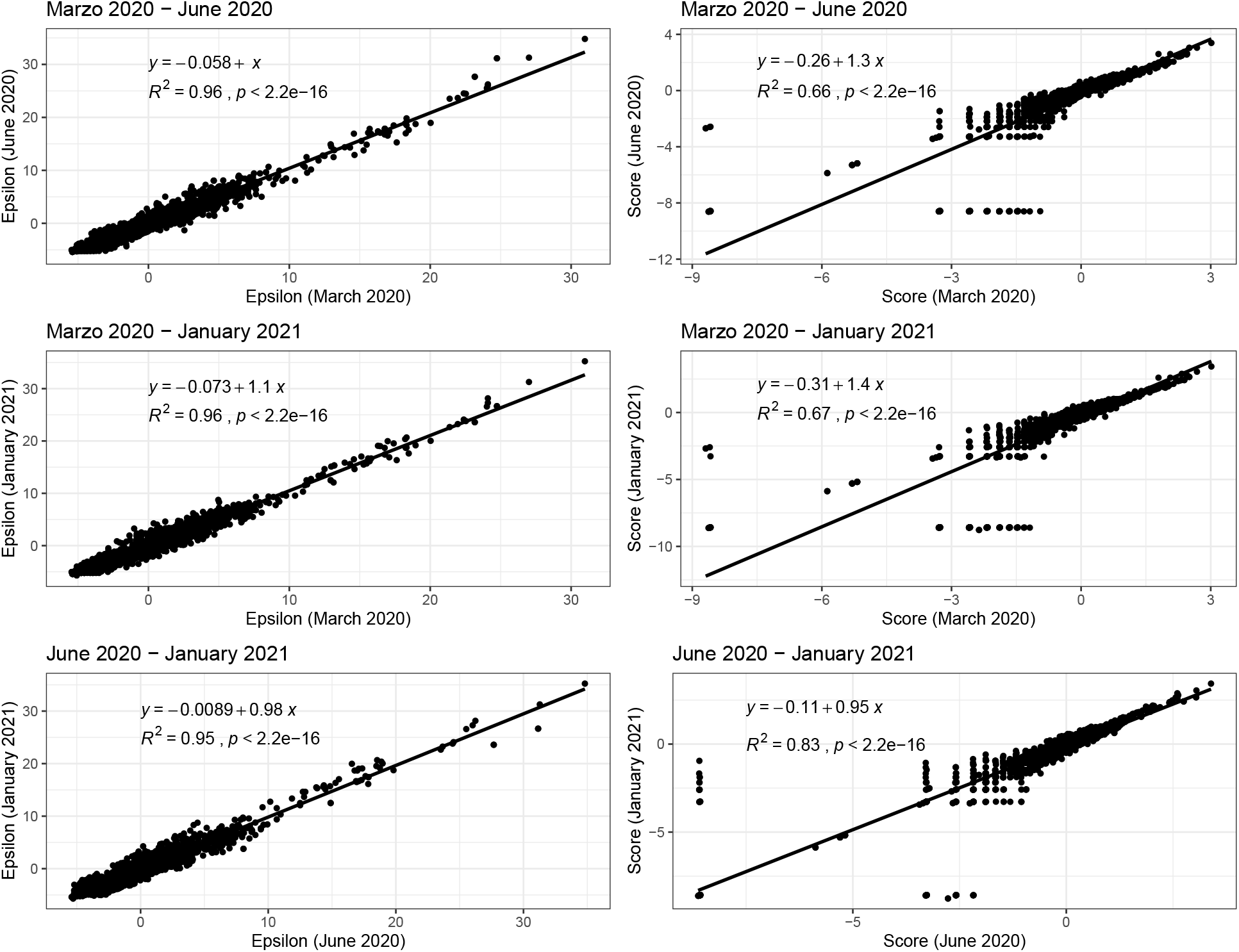
Left: Correlations between 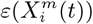 values for all habitat variables for the model with class top 10% of municipalities with highest number of cases for two different months; Right: Correlations between 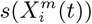 values for all habitat variables for the model with class - top 10% of municipalities with highest number of cases - for two different months.

In Figure 5 (left) we see the results for a purely spatial ENM/SDM model with target class, corresponding to question ii), being those municipalities with the presence of confirmed cases of COVID-19 for the months: March 2020, June 2020 and January 2021. Performance is again based on a 70%/30% train/test split of the 2458 municipalities, with Figure 5 (left) showing the corresponding ROC curves and the corresponding AUC for five separate SDM/ENMs for each test set, with: socio-demographic/economic variables only, mobility only, climate only, air contamination only and all factors together, for each of the three considered months. Again, the relative performance of each sub-model is clear, with the same relative performance as for the previous models. However, we note that the overall performance of the mobility, socio-demographics and All models is less than their counterparts in the case of top 10% of municipalities with most cases as class, while the performance of the climate and air contamination models is similar. In Figure 6 (left), in analogy with Figure 4, we see the correlations between 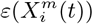 values for all habitat variables for the model for presence of COVID-19 cases for two different months, while in Figure 6 (right) we show the correlations between 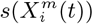 values for all habitat variables for the same model. Note that the correlation between the 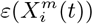 and 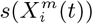 distributions in March 2020 and June 2020 and March 2020 and January 2021 are now significantly smaller than their counterparts for the top 10% model.

**Figure 5:**
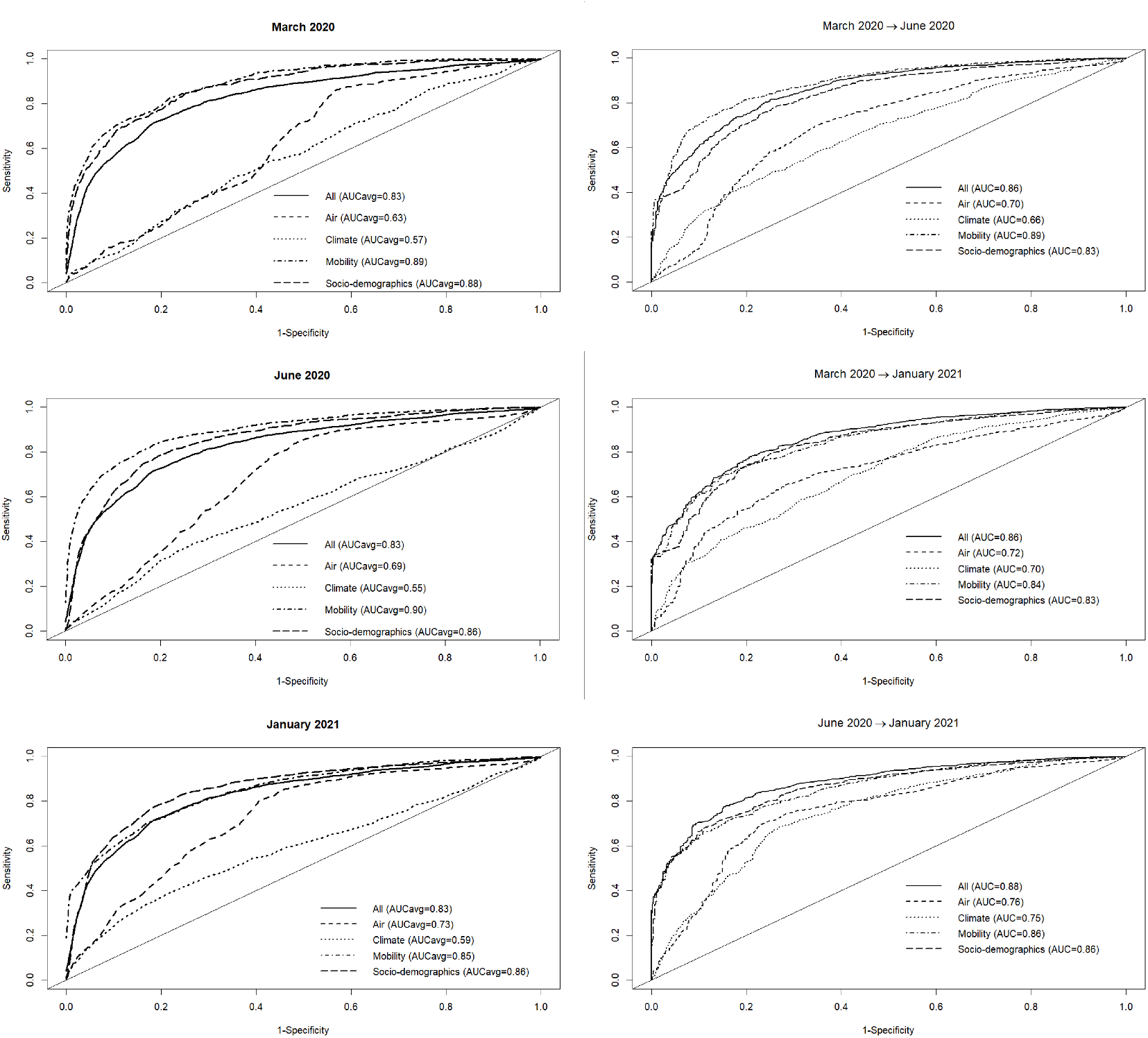
Left: Performance for 5 different SDMs for the class - presence of COVID-19 cases in the municipality - according to the habitat variable group considered (mobility, socio-demographics, air, climate and all) for three different months of the pandemic. Five different train-test 70%-30% splits were considered for each month; Right: Performance for 5 different SDMs for the class presence of COVID-19 cases in the municipality according to the habitat variable group considered (mobility, socio-demographics, air, climate and all) using a model trained on one month and tested on a future month.

**Figure 6:**
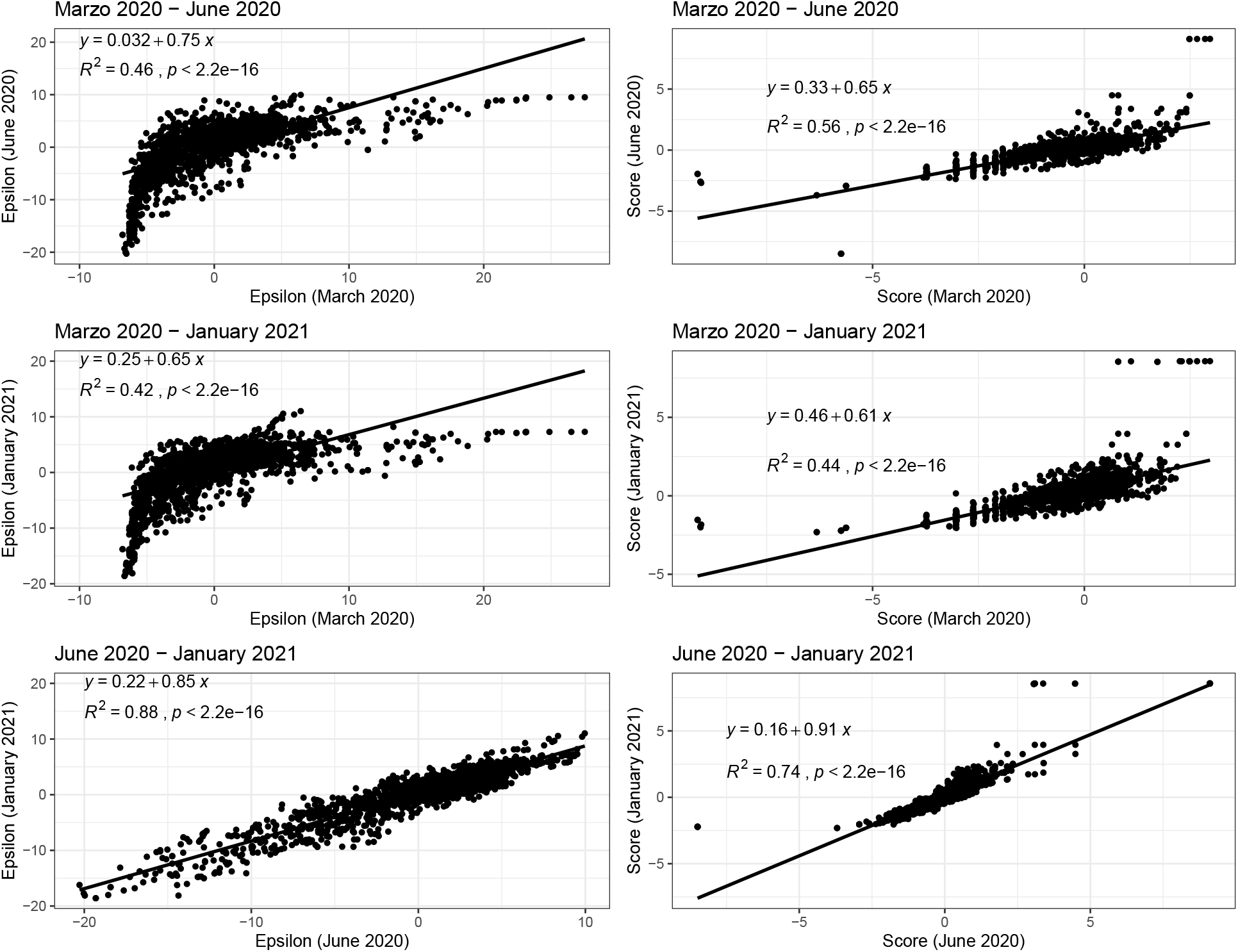
Left: Correlations between 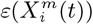 values for all habitat variables for the model for presence of COVID-19 cases for two different months; Right: Correlations between 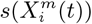 values for all habitat variables for the model for presence of COVID-19 cases for two different months.

Furthermore, in Figure 3 (Top) we show the values of the score 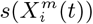 for the 10 deciles, *m* = [1, 10], of the habitat variable *X*_*i*_ = Internal labor flow of the municipality to compare and contrast the score contributions of this key niche variable as a function of time. We observe that the relative contributions from each decile differ significantly when compared to the results for The top 10% of municipalities with highest number of cases, as shown in Figure 3 (Bottom).

Turning now to the above ENMs at time *t* as predictors of the species distribution at *t*′, in Figure 5 (Right) we see the results of a model trained on data from those municipalities with confirmed cases in March 2020 and June 2020 to predict which municipalities would have cases in June 2020 and January 2021 and January 2021 respectively. In this case, we observe that the ENM for month *t* is less predictive for month *t*′ than the corresponding model for identifying the top 10% of municipalities with the highest number of cases.

### 2.3 ENMs can be used to predict numbers of cases

As well as using *S*(*C* | **X**) as a pure classifier (see Methods section), to identify a particular class, such as presence/absence, top 10% highest cases etc., we can also use it to predict the actual number of cases. To do this, as discussed in section 4.5, we regress the total score *S*(*C* | **X**) against the number of cases on our training set and then apply it to the test set. An example is shown in Figure 7, for the classification model with *C* = top 10% of municipalities with the highest number of cases, where the training set is a randomly chosen 70% of spatial cells and the test set the remaining 30%. All habitat variables were used. Figure 7 (left) shows the relation between score and number of confirmed cases for our three example months - March 2020, June 2020 and January 2021. First, municipalities were ranked according to their score, then grouped into 10 deciles. An exponential function fit was used. Figure 7 (right) shows the results of applying the model to the 30% out of sample test set, showing the relation between predicted and actual abundances.

**Figure 7:**
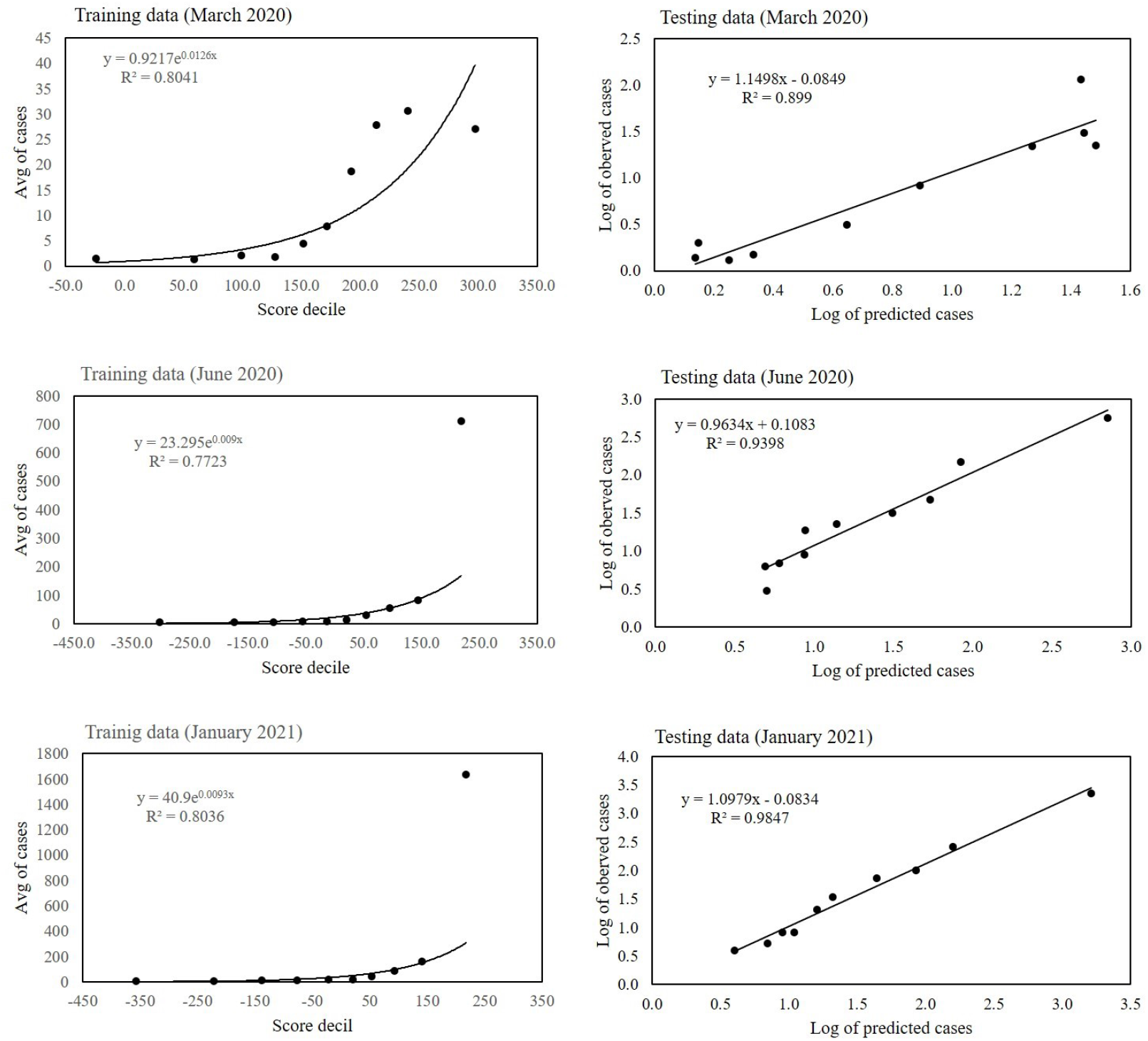
Left - graphs of score from a model for predicting the top 10% of highest number of cases versus number of confirmed cases of COVID-19 for the three months March 2020, June 2020 and January 2021 using as training set 70% of spatial cells. An exponential function was used to fit the relation score-number of cases. Right - graphs of number of predicted cases versus number of actual cases for the 30% hold out set for the same three months.

### 2.4 Causal relationships can be deduced

As an illustration of the formalism for disentangling causal relationships shown in section 4.6, we consider the relative contributions of climatic factors, as represented by the WorldClim variable *Average annual temperature*, and human mobility factors, as represented by *Internal labor flow of the municipality*, to prediction of the top 10% of municipalities with the greatest number of cases. The temperature variable is divided into 20 coarse-grained bins, while the mobility variable is divided into 10 bins. In any spatial cell, *α*, we may then determine if there is a presence or absence of a given coarse grained bin for either variable. Thus, in equation (4), we have 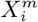 representing the 10 different bins of the mobility variable and 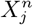 representing the 20 bins of the temperature variable. As, for a given spatial cell, there are four possibles states, corresponding to presence/absence of the two habitat variables, 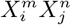 can be represented by four 20 *×* 10 matrices.

For each cell of each matrix we calculate *P* (*C* | *X*_*i*_*X*_*j*_) and also 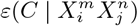, using as null hypothesis *P* (*C*), as shown in Figure 8, with data from January 2021.. Note that the absence of a variable bin, 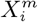, corresponds to those cells where there is no presence of 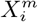. However, there will be a presence of at least one other 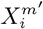, for *m*′ ≠ *m*. For example, in the matrix *Clim*0_*Mob*0, the cell *T mp.p*04 = 0, 08_*Inter* − *Mob* = 0 corresponds to those cells that are not in the average annual temperature range corresponding to *T mp.p*04 and also not in the Internal labor flow range corresponding to 08 *Inter* −*Mob*. However, that set of cells could have presences of temperature ranges other than *T mp.p*04 and mobility ranges other than 08_*Inter* − *Mob*.

**Figure 8:**
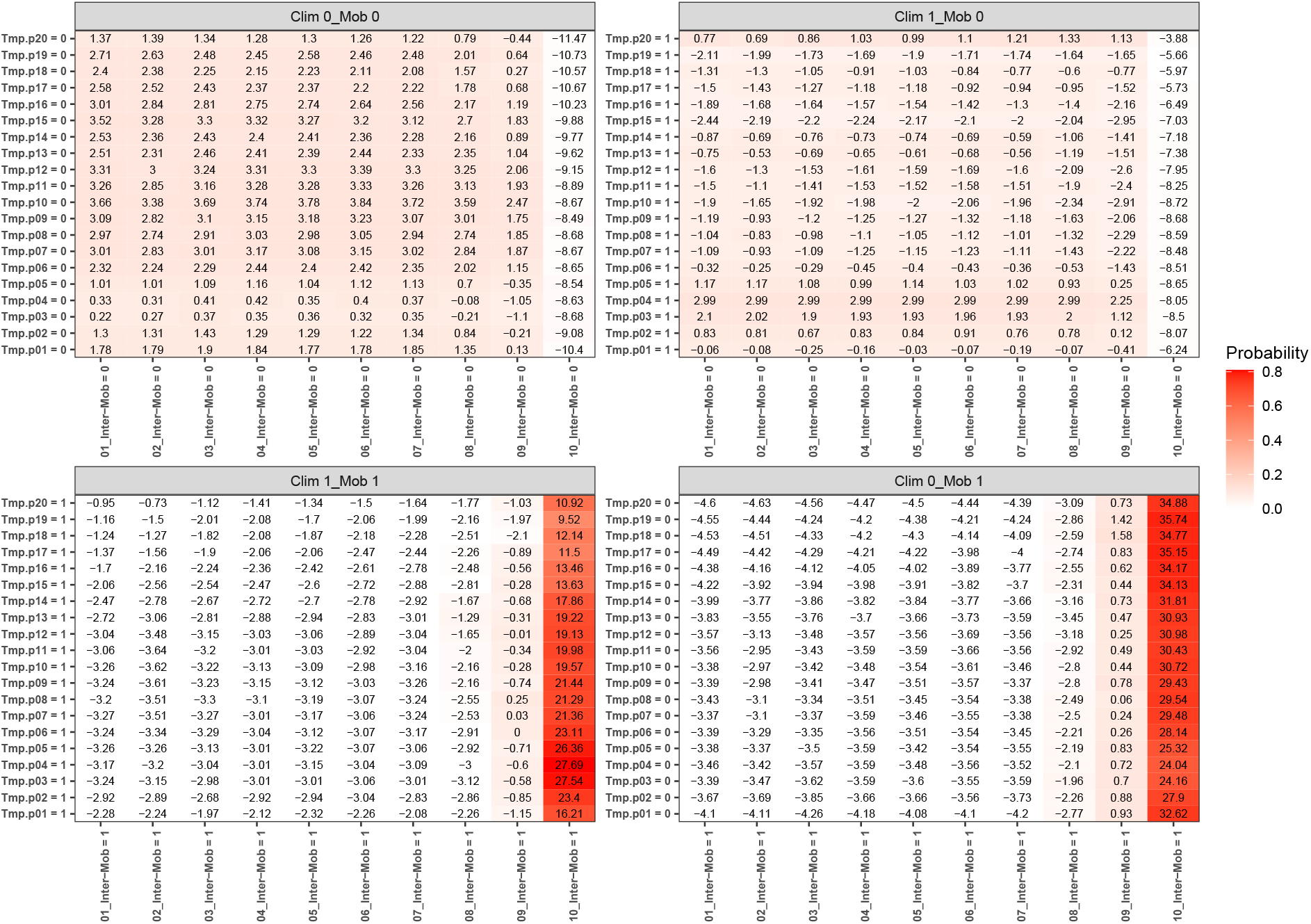
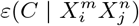 and 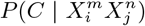 for each combination of habitat variable ranges 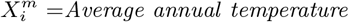 and 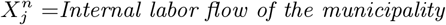 for the four combinations 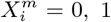 and 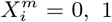 corresponding to absence/presence for each variable bin respectively.

The fact that the climate variable is confounded by the mobility variable is manifest in the matrix *Clim*1_*Mob*0, where there is a presence of a particular climate variable bin and an absence of the corresponding mobility variable bin. For instance, the cell *T mp.p*04 = 1, 10_*Inter* − *Mob* = 0 in that matrix corresponds to those municipalities that are not in the highest decile of mobility, but are in the fourth decile of average annual temperature. 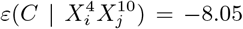 and 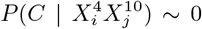 there, indicating that the probability of being in the top 10% of highest number of cases is very low. Indeed, we can see that for any temperature range, for 10_*Inter* − *Mob* = 0, there is very little chance of the corresponding municipalities being in the top 10%. On the contrary, for *Clim*0_*Mob*1, we see that 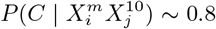 for any *m*, thus indicating that high mobility is very niche-like, independently of the value of the temperature variable. Although for *Clim*1_*Mob*0 the range *T mp.p*04 can give rise to statistically significant 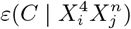 values for *m ≠* 10, by examining the case *Clim*1 *Mob*1 we see that the 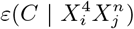 values for *n ≠* 9, 10 are all negative and statistically significant. Thus, climate may appear to be niche-like but, in fact, is confounded by mobility, and now we can intuit why a climate model can create predictability even though climate is not predictive in and of itself.

A model based on climate only is equivalent to the combination 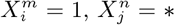, where *\** denotes that we have marginalized over this value. For instance, 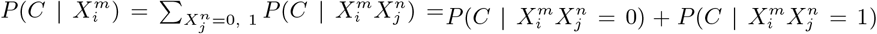. Although, 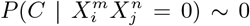 for *n ≠* 9, 10, 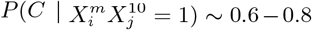 for any *m*. Thus, the primary reason why the climate model has some degree of predictability is that those places of highest mobility have a climate “profile”, i.e., they are not equally distributed across all average annual temperature ranges. It is not, however, the climate that is causal. Another way to see this is shown in Figure 9, where we show the marginal probabilities 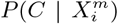 and 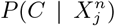, which would correspond to the results associated with an ENM based only on the climate variable *Average annual temperature* or on the human mobility variable *Internal labor flow of the municipality* respectively. We can note that, although the degree of predictability in the climate-only model is weak, there is a variation, with 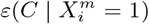 ranging from 0.9 to −2.9. However, the origin of the most niche-like value of 0.9 is associated with the residual predictability from decile 10 of the mobility variable.

**Figure 9:**
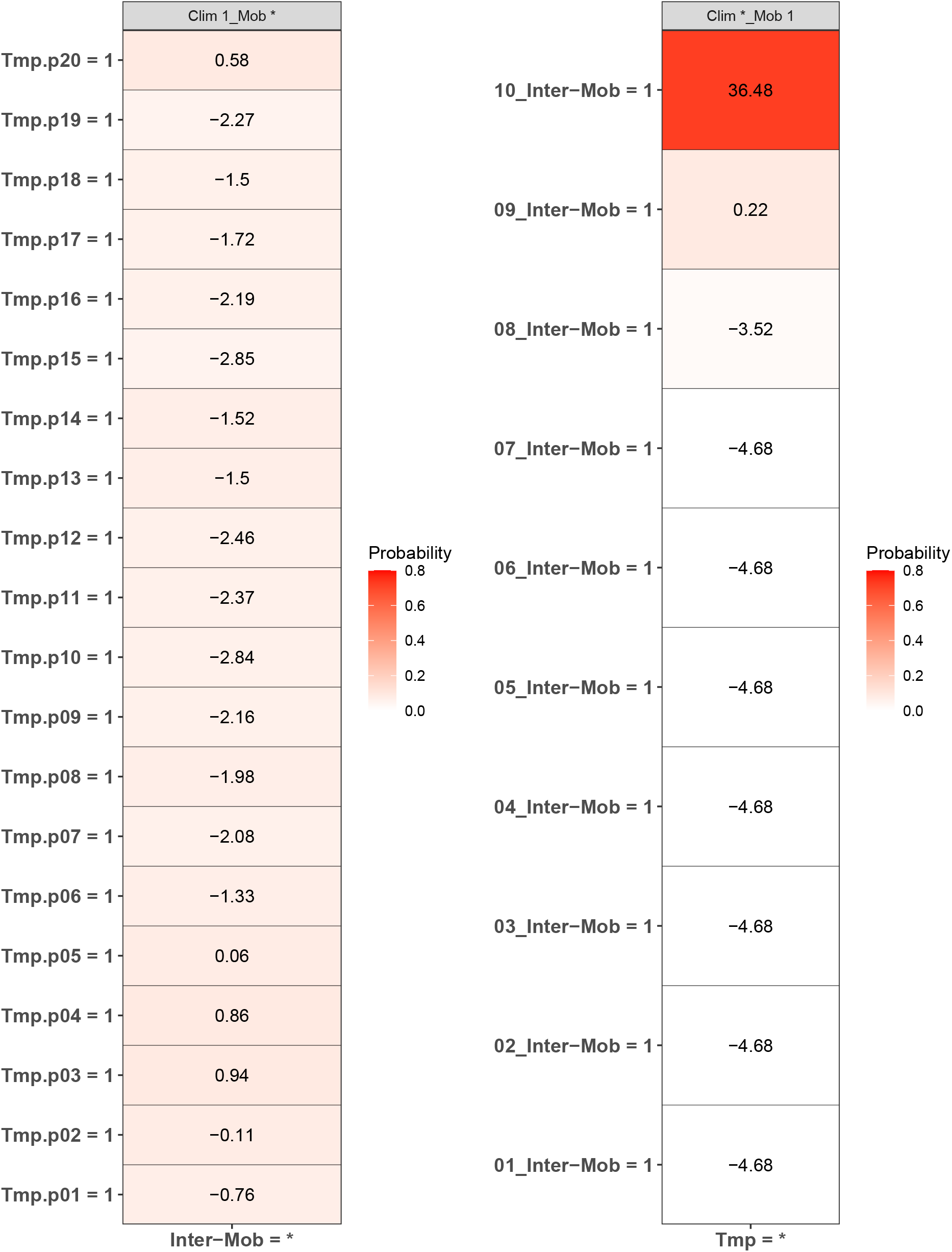
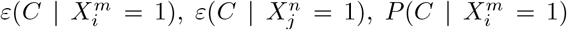 and 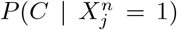 for each value of the habitat variable ranges 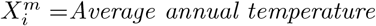 and 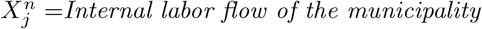 corresponding to a “climate-only” model and a “mobility-only” model.

## 3 Discussion

Our goal in this article was to demonstrate that the questions: “Does a respiratory virus have an ecological niche, and if so, can it be mapped?” can be answered in the affirmative. We have explicitly created several ENMs and SDMs for COVID-19 that are both predictive and contain habitat factors that are more causally plausible than climate, for instance. In order to do this we introduced several innovations compared to standard niche and species distribution modelling. Firstly, we showed how to extend niche and species distribution modelling to “non-equilibrium” situations, where both target and niche variables are potentially time varying, as well as the relation between them. Secondly, we created models with habitat variables that were represented by quite different data types and associated spatial resolutions. Finally, we showed how causal relations and confounding can be better understood by introducing a hierarchy of conditional probabilities and the associated intuition that a more causally direct factor should have a bigger effect than an indirect one.

We constructed ENMs as Niche Landscapes, *P* (*C*(*t*) | **X**(*t*)) - Bayesian posteriors which serve as a height function on a Hutchinsonian ecological space with 2749 dimensions, spanning air pollution, climate, mobility, socio-economic and socio-demographic data. Usually in ENM/SDMs, the target class, *C*, is a binary variable, such as presence/no presence of a species. In [5], the corresponding variable was “high” infection rate, defined in a binary fashion with respect to a reference infection rate. We too, have used binary class variables, by choosing a particular subgroup of spatial cells, corresponding to presence/absence of confirmed cases, or if a spatial cell was in the top 10% of highest total infections.^2^ However, the binary nature of the target variable is a choice rather than a restriction. For instance, taking infection rate as a continuous variable, this can be divided into as many quantiles as we please, say *n*_*c*_. The target variable now consists of *n*_*c*_ “presence/absence” variables, each with its own ENM/SDM, *P* (*C*_*i*_ | **X**), *i* ∈ [1, *n*_*c*_]. Even in the case of a binary decomposition of a continuous variable, however, the metric nature of the variable leaves an imprint on *P* (*C* | *S*(*C* | **X**)), such that higher score, as a continuous metric variable, corresponds to higher infection rate or number of cases, as we have demonstrated, leading to a model that can predict abundances.

In criticising ENM/SDMs as a useful tool for the COVID-19 pandemic, or any other, it is important to distinguish between applicability of ENM/SDMs in general versus a particular instance of an ENM/SDM. It is appropriate to criticise a model that includes climate, and which has been used to infer a corresponding causal effect on the pandemic, without any analysis of possible confounders. This does not mean, however, that with appropriate habitat variables and a methodology for disentangling confounding, that useful ENM/SDMs cannot be constructed, as we have shown here. Our results clearly show that models for predicting where the highest number of cases will be, that are built on mobility, socio-demographic and socio-economic data, are much more predictive than models built on climate and/or air contamination, as can be seen in the results of Figures 1, and that this has been true throughout the pandemic. As seen in Table 2, the most important niche/anti-niche factors for this target are all associated with the highest/lowest levels of mobility, as proxied by inter- and intra-municipal labor flows, and a particular socio-economic profile. The fact that a climate model can exhibit some degree of predictability does not mean that climate is the direct driver. On the contrary, we can ask if any apparent predictability due to climate factors is confounded by human-based factors. As we have shown in section 2.4, and is manifest in Figures 8 and 9, this is indeed the case. Just the distribution of the habitat variable scores themselves tells us that if there is confounding then it is the human factors that confound climate and not vice versa, as a confounder should be causally closer to its effect than the confounded variable and therefore have a higher score, as is implicit in the Bradford-Hill criteria.

With respect to the socio-demographic and socio-economic factors, we see that they have a similar predictive power to the mobility factors and reflect the socio-demographic and socio-economic conditions where COVID-19 cases are highest. For instance, the habitat variables *% of households that have a computer* and *% of households that have internet access* are both significant niche factors, as are other factors that correspond to a more educated population with higher economic status. Of course, the interpretation of these factors is not as intuitive as mobility and we certainly do not wish to attribute direct causality to them. However, there are a variety of relevant factors for COVID-19 that can be related to internet access and computer usage for example, such as age, educational status, population density [21], as urban areas have better infrastructure, and mobility itself [22, 23]. As with any epidemiological or ecological model, the interpretation of the predictors as representing direct versus indirect interactions is highly non-trivial. The formalism we have introduced for disentangling confounding can help in this regard.

The places where COVID-19 is in highest abundance represents one particular characterisation of the spatio-temporal distribution of the COVID-19 “species” and its associated ecological niche. Where it is present and where it will be in the future compared to where it is now are two others. Hutchinson [7, 24] defined niche as that region of ecological space where the net growth rate of the species is *r ≥* 0 at low density. For a pathogen that is not capable of free movement and is dependent on a host, there are two natural growth rates: one associated with the pathogenic load within a host and another that is measured by the number of infected hosts. Obviously, in this paper we are concerned with the latter, where the growth rate is characterised by the basic reproduction rate, *R*_0_, or the effective reproduction rate, *R*_*t*_ [25]. If we defined niche for COVID-19 through an analog of *r ≥* 0, such as *R*_*t*_ *>* 1, we would clearly be in a situation where we were passing from “niche” to “anti-niche” (*R*_*t*_ *>* 1 *→ R*_*t*_ *<* 1) and vice versa, continuously in space and time due to a multitude of factors, including public health interventions, such as lockdowns and vaccinations, as well as resistance to vaccinations, new variants and a host of others. That these factors alter the ecological conditions in a certain place at a certain time is undeniable. However, to keep track of such changes and how they impinge on how niche-like conditions are in a certain spatio-temporal cell, (*α, t*), requires the corresponding data. Here, we have preferred to use characteristics of the pathogen distribution that are easier to measure - number or presence of cases - but with which we can characterise the concept of niche, and a set of habitat variables that go beyond those previously considered. We take highest case abundance to distinguish those conditions in ecological space, and in geographical space and time, that favour higher abundance of the pathogen. This is a *relative* measure, as it may be that a municipality, *α*, has *R*_*t*_(*α*) *<* 1 but such that it is higher than any other.

Holt [26] has suggested that besides an “Establishment Niche”, that corresponds to the original Hutchinsonian niche, a “Population Persistence Niche”, associated with the range of niche space in which populations above some threshold density, *N >* 0, can persist, is a complementary notion. We take where COVID-19 is in highest abundance to be closer to this Population Persistence Niche, whereas where it is present is closer to the original Hutchinsonian characterisation and, especially, as it is portrayed in the majority of ENM/SDMs [27], where presence/absence is used to characterise both. On the other hand, where it is now versus in the past corresponds to neither, but is taken to reflect the potential range expansion/contraction of the species. We believe that these examples, and more, show that there are multiple characteristics of a species distribution that can and should be modelled and can be used to characterise complementary notions of niche.

The methods we have exhibited and the corresponding examples also allow us to understand to what degree a niche is conserved. In the case of species abundance, we saw that the niche associated with those places where the relative number of COVID-19 cases was highest was highly conserved, with very little difference across the entire pandemic. Moreover, we showed how this conservatism could be quantified using our statistical diagnostic 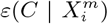 and the score functions, 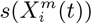, associated with the habitat variables 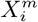, with the time dependence of the associated score function reflecting the relationship between target and habitat variables. For example, if 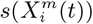 is strongly positive at one time, *t*, and not another, *t*′, then we may say that the variable 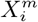 is niche-like with respect to *C* at time *t*, but not at *t*′. Niche conservatism with respect to 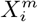 is then quantified by 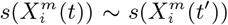. This niche conservatism is also manifest in that an ENM created at time *t* provides a SDM that is just as predictive at a later time *t*′ as at *t*. Thus, from a Hutchinsonian perspective, the relation *P* (*C*(*t*) | **X**) may be conserved in ecological space, even though the spatial distribution of *C*(*α, t*) and/or **X**(*α, t*) could change in time.

In the case of presence/absence as target class, the corresponding realised niche is not conserved, in that 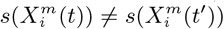. This is seen in Figure 6, when compared to Figure 4, where the regression slopes for the March 2020-June 2020 and March 2020-January 2021 comparisons have lower *R*^2^ values and also slopes *<* 1, indicating that the scores in January or June are only about 60% of their values in March. The *R*^2^ values and slopes for the *ε* comparisons in the same figures show the same effect. These differences just reflect the range expansion of COVID-19, where it has been argued that: “there is no unsuitable habitat” for COVID-19 [2]. This is linked, however, to the notion of presence, not to abundance. The fact that the score and *ε* contributions are diminishing in the presence model from March 2020 to June 2020 and January 2021 is due to the fact that the habitat variables are less able to discriminate between those cells where cases are present versus absent. The differences between June 2020 and January 2021 are much less as, by this time, a large majority of municipalities now had confirmed cases. We can also see this niche non-conservation in Figure 4 (top), using as an example the variable *Internal labor flow of the municipality*. We see that in March 2020 only the first three deciles of municipalities ranked by that score are associated with a positive score - niche-like - whereas in June 2020 and January 2021 60% have positive scores. Similarly, we see that in the most anti-niche-like deciles, *Decile* 02 and *Decile* 01, the scores are becoming less negative, indicating that those municipalities with the lowest internal labor flows are becoming less anti-niche-like. If the pandemic had a presence in *every* municipality, then every 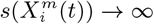, corresponding to the fact that there can be no discrimination between where the species is present and where it is absent. Everywhere is niche-like. This would not be the case, however, for abundance, as is seen in Figure 4 (bottom), where the change is due to the fact that our relative abundance measure is associated with the top 10% of municipalities with the highest number of cases. The range expansion of COVID-19 presence also has an impact on the performance of the corresponding SDMs, as seen in Figure 5, where we see that for the All, socio-demographic and mobility models, the corresponding AUC for the presence/absence spatial models is much less than their abundance counterparts.

We have also shown how a classification-based ENM can be used to predict abundances, with an example being the number of confirmed cases of COVID-19 for a given month. We see that the score based on all habitat variables explains approximately 90% of the variance. We believe that the ENM/SDM formalism we have developed here has the capacity to be truly epi-ecological/eco-epidemiological given the right habitat variables. The static habitat variables we have chosen cannot account for the dynamic expansion and contraction that is characteristic of epidemics and which naturally emerges from a mechanistic SIR-type modelling formalism. What is required are habitat variables that are the analog of what enters into a differential equations type modelling environment: changes in abundances from one time period to another, for example, or even changes in those changes, as it it these variables that account for the underlying dynamics. Indeed, an ENM built on a given time slice that does not explicitly account for such variables will either underestimate or overestimate abundances, depending on whether the epidemic is in an expansion or contraction phase. We will return to this in a future publication.

We have also shown how it is possible to distinguish confounding and how this can be used as a tool to disentangle causality. As a test case we considered combining a behavioural/socio-economic/socio-demographic variable - *Internal labor flow of the municipality* - and a climatic variable, showing how climate as an apparently predictive habitat variable is confounded by the more predictive socio-economic and mobility factors.

An apparent limitation of our work is that we have worked at a quite coarse-grained level, that of municipalities. This is a limitation of the data used, however, not the methodology. In principal, a much finer spatial resolution, at the level of “census blocks”, for instance, could be used if data was available at that resolution for the target class and habitat. In the same vein, dynamical data that represented both changes in the local environment, such as lockdowns and hospital occupation rates, or changes in behaviour, such as mask wearing compliance, cell phone data as a proxy for short-term mobility, or vaccination rates, or a host of other factors could also be included. It would be interesting for example to determine to what extent adaptations in the pathogen were potentially reflected as changes in its niche. There is also the question of the validity of the target class data as maintaining accurate counts of confirmed cases is difficult. However, this would have little to no impact on the overall conclusions of our ENM/SDMs.

In summary: Our goal here was not to offer a gold-standard model for prediction of the pandemic. As mentioned, there are more predictive and directly causal factors than we have included here. However, the niche of COVID-19 is immensely complex and adaptive and the incorporation of the vast array of relevant factors that determine it is a huge challenge in data collection and integration. What we have shown is that if that data can be represented in space and time, **X**(*α, t*), then it may be incorporated into an ENM/SDM using the methodology we have shown here. Moreover, the innovations we have shown are independent of the specific use case of COVID-19 considered here. They represent general extensions of current niche and species distribution modelling, and can be applied to any ecological system, where they are necessary. In particular, they can be applied to situations where the target and/or niche variables are changing in time. Besides disease dynamics, invasive species and habitat and biodiversity loss are other prime areas where time dependence is crucial and where our approach can be used. Moreover, we believe that taken in its fullest sense, where a niche/anti-niche represents the complete set of both biotic and abiotic drivers that favour where a “species” is or isn’t is, or at least should be, a universally applicable concept, with the SDM determining the “where” and the ENM the corresponding “why”. Indeed, its applicability is only restricted in an ecological setting by just what we mean by “ecological”. If we take ecology to cover any interaction between biota and the environment then we should accept factors as mask-wearing compliance as a potential niche factor for example. Furthermore, when thinking of a “Species” Distribution Model, we may encouraged to be liberal in our notion of “species”, where it may represent, for instance, cases of non-transmisable diseases, such as diabetes or heart disease.

In conclusion, there is a difference between stating that ENM/SDMs generally are inappropriate vehicles for modelling a dynamic phenomenon such as the *COV ID −* 19 pandemic versus stating that a particular ENM/SDM is inappropriate. We have shown that ENM/SDMs can be generated which overcome such criticisms.

## 4 Methods

Several of the methodological elements used in this paper have been used previously for creating ENM/SDMs in multiple contexts [28, 29, 30, 31]. Further details can be found in these papers and in the Supplementary Material.

### 4.1 Defining a spatial grid

All SDM/ENMs are based on the notion of co-occurrence between a target, *C*, and, one or more, predictors/habitat variables, **X** = (*X*_1_, *X*_2_, …, *X*_*N*_). Normally, co-occurrence is considered purely in spatial terms, although the concept can be extended to co-occurrences in time. In either case, to specify whether there is a co-occurrence or not requires a specification of a grid that divides a spatio-temporal region into cells. A spatial grid can consist of cells of arbitrary shape, as long as they form a partition; i.e., each spatial point is a member of one and only one cell. A partition may be uniform, such as formed by rectangular cells of a given area, or irregular, as is the case for political/administrative units, such as municipalities, counties, states etc. In standard SDM/ENM modelling [27] this partition is implicit, corresponding, for example, to the pixels of environmental rasters. Such a partition, however, creates a barrier when wishing to include biotic factors, such as point collection data, that cannot be naturally represented as a raster. In short, we may wish to ask: what is the relative importance of average annual temperature, as taken from WorldClim, versus the presence of a prey species in an SDM/ENM for a vagile carnivore?

In [32] a methodology has been developed for incorporating spatial data layers of arbitrary type and spatial resolution. A spatial grid is overlaid on a chosen spatial region and co-occurrences defined with respect to a given cell. Thus, if there is a co-occurrence between *C* and a particular variable, *X*_*i*_, in a particular cell, then *N* (*CX*_*i*_) = 1. In the case of a continuous variable, such as species abundance, temperature or precipitation, the variable, *X*_*i*_, is coarse grained into a set of *n* discrete bins, leading to *n* discrete “presence/absence” variables. Thus, 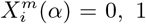 represents the presence/absence in a cell, *α*, of values of *X*_*i*_ in the range defined by the *m*th bin. Any categorical variable can be left as is, or also coarse grained into a smaller number of categories, if necessary. The criterion for fixing the bin distribution of a variable are that it allows for the best discrimination - dependence of *C* on *X*_*i*_ - while maintaining an appropriate degree of statistical significance - number of cells associated with a given 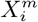. Furthermore, the target *C* can also be discretised if necessary in the same way, with bins 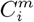. In summary, all variables become categorical, with each category being associated with a binary variable.

An advantage of this categorisation is that no relation is assumed between one bin and another, as would be the case in a regression-based approach for example. Using a non-uniform grid however, can introduce some bias, as some municipalities are bigger than others. We have, however, used spatially uniform grids without substantial changes in our results.

### 4.2 Constructing SDMs and ENMs

Having transformed all variables to this binomial form, counts, 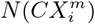, can be made over the region of interest, corresponding to the number of cells that contain a presence of the target 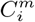 and the variable 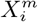. The number of cells associated with a given 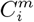 and/or 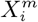 may be fixed or varying depending on the target class. For instance, if we define the class using a relative measure, such as the 10% of cells with the highest abundance for a given species, or the highest number of confirmed cases or deaths in the case of COVID-19, then the class will always be associated with 10% of the cells, independently of time. However, if the class is based on an absolute measure, such as presence/absence of the species, or confirmed cases of a disease, the number of cells in the class may vary in time as, for example, in the case of an invasive species, or an emerging epidemic. Similar considerations hold for 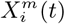. If it represents a relative measure, such as the 10% of cells with highest average annual temperature, then it will always cover 10% of cells. However, if it represents the presence/absence of an invasive species, the number of associated cells will change. We will consider these cases in more detail below.

From the co-occurrence counts, probability distributions may be calculated, such as 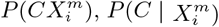 and 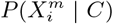, which are related through Bayes theorem. These distributions can be compared to a null hypothesis and a binomial test used, for instance, to determine the statistical significance of the deviation from this hypothesis. For example, for the posterior, a natural null hypothesis is *P* (*C*) ^3^ and a statistical diagnostic for determining if the habitat variable 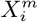 is significant or not is

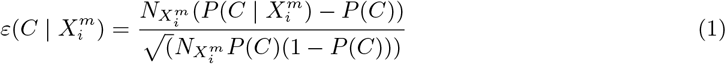

In the case that the binomial distribution can be approximated by a normal distribution, 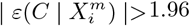 is equivalent to the 95% confidence interval. For a multivariate niche the corresponding distributions of interest are: *P* (*C***X**), *P* (*C* | **X**) and *P* (**X** | *C*). Although these exist formally from a frequentist perspective, i.e., *P* (*C* | **X**) = *N* (*C***X**)*/N* (**X**), in the case of a high-dimensional habitat, both *N* (*C***X**) and *N* (**X**) = 0, 1, which means that a direct statistical estimation is impossible. To overcome this problem, in [34], the likelihood *P* (**X** | *C*) has been estimated by assuming a factorisation of the form

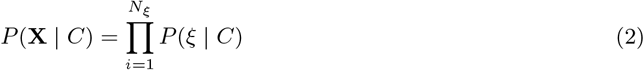

where *ξ* is a combination of a small number of variables. Thus, the abiotic and biotic habitat variables are partitioned into a set of *N*_*ξ*_ non-overlapping combinations. In the case that *N*_*ξ*_ = *N*, this corresponds to the well known Naive Bayes approximation [35]^4^ Usually, in order to calculate *P* (*C* | **X**), as the evidence function *P* (**X**) is independent of *C*, to omit it, the following “score” function is often used

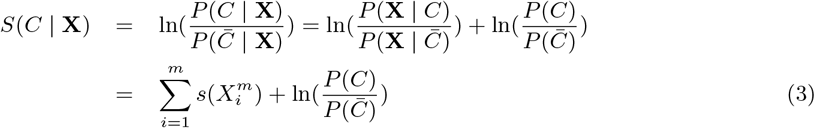

where 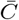 is the set complement of *C*, with 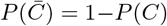, and 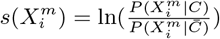 is the contribution (“score”) to the overall *S*(*C* | **X**) from the variable 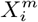. If 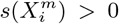, *<* 0 then the factor 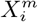 contributes positively/negatively to the occurrence of *C*. Everything now is based on simple cell counts: 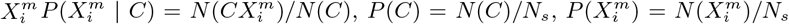 where *N* (*C*) is the number of cells with a presence of the target 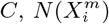 is the number of cells with a presence of the variable 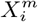 and *N*_*s*_ is the total number of cells in the spatial grid. *P* (*C* | **X**) can be determined directly from *S*(*C* | **X**) by deriving the relation *P* (*C* | *S*(*C* | **X**)) = *N* (*CS*)*/N* (*S*), discretizing the score range into bins and then counting how many cells in a given score range also have a presence of the target. Δ(*C*, **X**) = (*P* (*C* | **X**) *− P* (*C*)) is a measure of how niche-like the conditions specified by **X** are. The most niche-like conditions, **X**_*n*_, are those where *P* (*C* | **X**) reaches its maximum value, while we can term those conditions, **X**_*an*_, where *P* (*C* | **X**) reaches its minimum value, as the most “anti-niche” like. *P* (*C* | **X**) represents a height function for a given point in an *N* -dimensional Hutchinsonian ecological niche space. The corresponding “Niche Landscape” is our ENM. As every spatial cell *α* can be assigned a corresponding niche profile, **X**(*α*), *P* (*C* | **X**(*α*)) for different spatial cells also now yields an SDM over our spatial region of interest.

A significant advantage of assuming a factorisation of the likelihood is that the subsequent model is completely transparent, with each factor contributing separately, so that each factor can be compared and contrasted with the rest. Moreover, the same applies for groups of factors, so we can compare the relative predictive and discriminative value of climate factors versus socio-economic factors, or air contamination versus mobility.

Of fundamental importance in the construction of *P* (*C* | **X**) is the statistical ensemble from which the counts will be made. Although we consider a spatial ensemble, the data assigned to each cell invariably has a temporal dimension. In the case of standard SDM/ENM, with a target class defined through point collection data, each data point, *d*(**x**, *t*), is associated with a spatial specification, usually latitude and longitude, and a collection date, *t*. Similarly, abiotic factors are also time-stamped. A model for *P* (*C* | **X**) assumes that the distributions in time of both *C* and **X** are both statistically constant and representative.

In the case that the target variable is metric, such as number of cases, *N*_*C*_, the classifier *S*(*C* | **X**) can also be used to predict *N*_*C*_ (*α, t*) for a given spatial cell and over a particular time period. In the case of a spatial model, the cells may be divided up into training and test sets. The points (*N*_*C*_ (*α*), *S*(*C* | **X**(*α*))) for each cell, *α*, of the training set can then be plotted, and a regression performed to determine the relationship *N*_*C*_ = *F* (*S*(*C* | **X**)). This relation can now be used to predict *N*_*C*_ for any cell in the test set given we have *S*(*C* | **X**) there.

### 4.3 Dynamical ENMs and SDMs

A key aspect of the COVID-19 pandemic is its highly dynamic nature, which can manifest itself in several different ways. First of all, the habitat variables themselves may be time dependent, *X*_*i*_ *≡ X*_*i*_(*t*). Secondly, the target itself may be dynamical, *C ≡ C*(*t*). In the latter case, this may represent the fact that a disease or a species/disease is present at a given spatial point at one point in time but not another. Thus, the case of an invasive species would fit into this category. In the former, the disappearance of food resources or climate change would naturally fit. In general, we wish to model *P* (*C*(*t*) | **X**(*t*′)). It is important to point out that there is a difference, however, between having a ENM that is dynamical versus just considering a different configuration of the habitat variables substituted into a given model. In the case of climate change, for instance, this is usually done [36] by determining a static ENM, equivalent to *P* (*C* | **X**(*t*)), which can be applied to any spatial point, *α*, then using a climate change model to determine **X**(*α, t*) *→ X*(*α, t*′) for some future time *t*′. In other words, we assume the ENM doesn’t change, only the spatial distribution of the habitat variables, which is then used to determine the new spatial distribution of the target with the same original model. In other words, we determine an SDM at *t*′ using an ENM derived at time *t*.

To go beyond this limited context, we must return to the question of spatio-temporal cells as opposed to just a spatial grid. Considering a timespan *T*, we divide *T* into *N*_*T*_ intervals. We now have a total of *N*_*Ts*_ = *N*_*s*_ *× N*_*T*_ spatio-temporal cells. The simplest division is into uniform intervals, such as a year into 12 months. In a static setting, the interpretation of *P* (*C* | **X**) is that it represents an equilibrium relation between *C* and **X**, even though the data used to calculate it, more often than not, spans substantial and, effectively, non-commensurate time periods. For example, using WorldClim data from a given year as habitat variables for a niche model of a species represented by collection data taken from a 150 year time period. Without assuming equilibrium we should calculate *P* (*C*(*t*) **X**(*t*)). However, we may also consider calculating *P* (*C*(*t*) |**X**(*t*′)), i.e., to predict the effect of the habitat variables **X**(*t*′) at time *t*′ *< t* on our target *C*(*t*). Of course, an important question at the heart of this is: how are changes in **X**(*t*′) reflected in *C*(*t*)? This requires longitudinal observations and/or an understanding of the underlying causal relationships between the *X*_*i*_ and *C*.

There are four distinct paths to incorporating time into the SDM/ENM: i) assume equilibrium, and thereby ignore time dependence; ii) construct a model *P* (*C*(*t*) | **X**(*t*)) using a time slice/history to predict a spatial distribution *C*(*α, t*) on that time slice, as a function of the habitat variables on the same time slice; iii) predict in time, assuming niche conservatism, i.e., construct *P* (*C*(*t*) | **X**(*t*)) and use it to predict the spatial distribution, *C*(*α, t*′) at some later time using the habitat **X**(*α, t*′); iv) predict in time, without assuming niche conservatism, by constructing *P* (*C*(*t*) | **X**(*t*′)), where *t*′ *< t*. Models of type i) and ii), we term *spatial prediction* models. They are analogous to standard SDM/ENMs, in that they predict a spatial distribution. However, in distinction, in case ii) they do so using a time slice of data that permits us to compare and contrast *C*(*t*), **X**(*t*) and *P* (*C*(*t*) | **X**(*t*)) over time. For instance, we may use data only from May 2021 and compare with data from May 2020, or we may consider data from all of 2021 etc. Models of type iii) and iv) we term *time prediction* models. In this case they are an ENM/SDM equivalent to a SIR-type model, predicting the evolution of “where” in time, as opposed to “who”. The case with niche conservatism would be equivalent to a SIR type model, where the parameters of the model do not change, whereas the non-conservative case would correspond to the case where they do change and are fitted to dynamic data.

The notion of a *time prediction* model also naturally leads us to consider ENM/SDMs that are associated with changes in the distribution of a species/disease. For instance, we may consider the set of cells, *C*(*t −* 1), that have a presence in the time interval *t −* 1 and the set of cells, *C*(*t*), that have a presence in the time interval *t*. Δ_*C*_ (*t, t −* 1) may then represent those cells that had a presence/absence at *t −* 1 and, in contrast, an absence/presence at *t*. This could, for example, model the range expansion of a species. In this case, an ENM *P* (Δ_*C*_ (*t, t −* 1) | **X**(*t −* 1)) represents a model for predicting changes in the distribution due to the habitat variables. This model can then be applied to produce an SDM *P* (Δ_*C*_ (*α, t* + 1, *t*) | **X**(*α, t*)) using the habitat variables at *t* and predicting the change in distribution between *t* and *t* + 1.

We give more details about how to construct the different spatio-temporal models in the Supplementary Material 4.7. As discussed above, spatially, we used a grid corresponding to the municipalities of Mexico. For the temporal partition, we chose a month, though any other timescale of interest could have been used.

### 4.4 Testing model performance

The way in which our models are tested depends on the type of statistical ensemble that is used to train and then test the model. In the case of an ENM on a given time slice, *P* (*C*(*t*) | **X**(*t*), the model will be created on a training set that corresponds to a randomly chosen fraction, *f*_*train*_, of spatial cells with data associated with a time period *t*. The model is then tested on the remaining fraction of cells *f*_*test*_ = 1 *− f*_*train*_ from the same time period. The test and train sets can be chosen in multiple ways. Here we consider a 70%/30% split. Standard performance statistics can then be determined, such as from a confusion matrix or from the area under the ROC, among others [37, 38]. However, we can also use the ENM *P* (*C*(*t*) | **X**(*t*) trained on 100% of spatial cells and apply it to a future time period *t*′. In this case, the entire set of spatial cells at *t*′ is the test set. The luxury of a problem such as COVID-19 is that we have relatively accurate data about the spatial distribution of the disease as a function of time. In this case, the ENM *P* (*C*(*t*) | **X**(*t*) is used by substituting at *t*′, **X**(*α, t*′) for each spatial cell, *α*, to obtain for that cell *P* (*C*(*α, t*′) | **X**(*α, t*). We can now test the quality of the prediction using any of the above standard metrics, given that *P* (*C*(*α, t*) | **X**(*α, t*′) is a classifier, and we can compare with the actual distribution *C*(*α, t*′). We will expect the time *t* ENM to yield good performance at *t*′ only if the niche is conserved over this time period.

Besides the pure assumption of niche conservatism, where we apply the ENM *P* (*C*(*t*) | **X**(*t*)), we can compare and contrast ENMs from different time slices to determine the degree to which they are changing. This would correspond to determining if the niche is actually changing over time. This can be done for each habitat variable by comparing 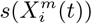 with 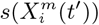. If the niche is changing then we must consider just how fast it is changing. This can be deduced by comparing the 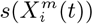 across time. If 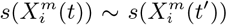 for two time periods *t* and *t*′ *> t*, then the niche is conserved over the interval (*t*′ *− t*). If they differ substantially, then we may reduce (*t*′ *− t*) and determine how much change there has been over this shorter interval.

For time prediction models that do not assume niche conservatism, the training set consists of choosing two time periods *t −* 1 and *t*. An ENM *P* (*C*(*t*) | **X**(*t −* 1)) is created on all spatial cells. This model is then applied to all spatial cells for the time periods *t* and *t* + 1 as test set. Thus, we use the ENM *P* (*C*(*t*) | **X**(*t −* 1)), substituting **X**(*α, t*) to create the SDM *P* (*C*(*α, t* + 1) | **X**(*α, t*)) for prediction of *C*(*α, t* + 1). We can then apply any of the above standard metrics to evaluate the performance of this ENM/SDM.

### 4.5 Predicting abundances

ENMs that are based on presence/no presence are, naturally, binary classification models. Although a metric variable, such as abundance, can be treated as such by considering multiple classes, it is also possible to use a binary prediction model, such as the top 10% of municipalities with highest confirmed cases, to construct a relation, *N* (*S*(*C* | **X**)) between the score, *S*(*C* | **X**), and the number of cases on a training set. This model can then be applied to a test set, predicting the expected number of cases for a given cell, *α*, using its habitat profile **X**(*α*). This procedure can be used for both *spatial* and *time* prediction models. In the former case the abundance predictions are associated with a given time slice *t* using a split of the cells into training and test sets, while for the latter we predict from a training set that consists of all cells on a time slice *t* to predict abundances on a test set of all cells on a time slice *t*′.

### 4.6 Inferring causality in ENMs

Another criticism of the application of standard SDM/ENMs to the pandemia has been the lack of plausibility of the relation between, say, infection rates and habitat variables, such as climate or contamination. As pointed out in [34], this is a problem with correlative approaches in general. This criticism however, can be applied to a phenomenological study of any Complex Adaptive System, where cause-effect relations are multi-layered and, therefore, often indirect. Epidemiology and medicine in general are rife with problems of causal inference and there are two basic approaches to it: a “classic” approach [39] and a “modern” approach [41, 42, 40]. An important criterion from our perspective is that of “strength of association” [39], where, although a small association does not mean that there is no causal effect, the larger the association, the more likely that it is causal. This is key to our use and understanding of both 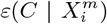 and 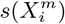 and their multi-factorial counterparts *ε*(*C* | **X**) and *s*(**X**). Thus, for two niche factors, *X*_*α*_ and *X*_*β*_, if 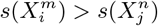, then we will judge 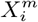 to be causally closer to *C* than 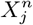. An illustrative example of this would be a food chain: carnivore *→* herbivore *→* plant food *→* climate, as considered in [34].

In [34, 43] we have proposed a formalism for examining causality that is particularly natural for application to ENMs. If we have two niche factors 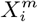 and 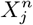, we can better understand their potential causal relations with a target *C* by considering the following relations

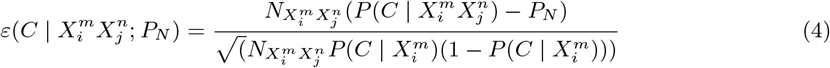

where *P*_*N*_ represents the null hypothesis with respect to which we will determine the predictability of the combination 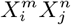. If *P*_*N*_ = *P* (*C*) we are gauging the consistency of 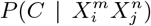 with the null hypothesis that the distribution of the target species is independent of the variable combination 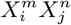. However, we may also choose as null hypotheses 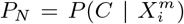 or 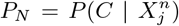, in which case the null hypothesis is that 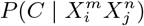 is independent of 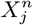 and 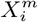 respectively. With this approach we can determine the degree to which 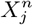 confounds 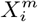 or vice versa. For example, if 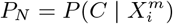and 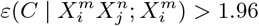 we can conclude that within a 95% confidence interval that 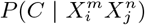 is not consistent with the null hypothesis and therefore the habitat variable 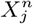 is predictive of the distribution of *C* beyond what is explained by the habitat variable 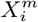. As 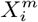 may be a biotic variable and 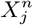 an abiotic one, we have used this to show that, very often, biotic factors are confounders for abiotic factors and not vice versa [34]. Although, here, we are concentrating on causal relations and confounding with respect to pairs of habitat variables the formalism naturally extends to larger numbers of variables.

In any spatial cell *α* we may determine if there is a presence or absence of a given coarse grained bin for either variable, 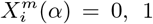 and 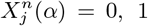. For example, 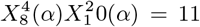 would represent a presence of bin 4 of variable 8 and a presence of bin 2 of variable 10 in the cell *α*, while 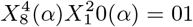 would represent an absence of bin 4 of variable 8 and a presence of bin 2 of variable 10 in the cell. Thus, there are 4 possible combinations for any pair of habitat variables: presence-presence = 11, presence-absence = 10, absence-presence = 01, absence-absence = 00. By comparing and contrasting the different combinations we may determine, for example, if presence of one habitat variable is more predictive than the other. Note 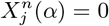 does not imply absence of the variable itself in the cell, just the range denoted by the *n*th coarse grained bin.

### 4.7 Data and habitat variables

In this paper, considering the evolution of the pandemia in Mexico, we used an irregular spatial partition consisting of the 2458 municipalities of Mexico. The advantage of this partition is that it is most aligned with publicly available socio-demographic and socio-economic factors that can serve as corresponding niche variables. It’s disadvantage is that there is a potential bias in the spatial distribution of different municipalities according to their area. For the epidemiological data, used to define the target classes, we used data from the Direccíon General de Epidemiología de la Secretaria de Salud [44]. For the socio-demographic and socio-economic data we used 124 variables taken from the 2010 Mexican Census [45] at the municipality level. For mobility data we used 12 variables provided by the Institute of Geography of UNAM, that represent the average, daily labor flows between a pair of municipalities. For the air contamination factors we used three atmospheric pollutants (Formaldehyde (HCHO) Nitrogen dioxide (NO2), Sulfur dioxide (SO2))[46, 47, 48], while for climactic data we used 19 bioclimatic variables from the WorldClim database (www.worldclim.org) with a spatial resolution of 30 arc-seconds (*≈*1km) [49], which includes 11 temperature and eight precipitation indices that express annual trends (e.g., annual mean temperature and precipitation), seasonality (e.g., annual temperature and precipitation ranges), and environmental extremes (e.g., highest and lowest values of temperature for the warmest and coolest months).

As all the socio-demographic, socio-economic and mobility data was metric and continuous, for each variable we ranked the 2458 municipalities from highest to lowest value then divided the ranked list into deciles, with 10% of all municipalities in each decile. By doing this, as opposed to dividing the metric interval for the variable into equal parts, we assure equal statistical weight to each coarse grained category. For the air pollution variables we divided each raster layer into 20 ranges, which were chosen to have roughly equal number of pixels in each one. Climate variables were also divided into 20 ranges. Thus, in this way we mapped both target class and habitat variables in their entirety into a set of binomial presence/absence variables for each cell on our grid. We also included in as a habitat variable the decile of confirmed cases associated with a given municipality, but at period *t −* 1, as opposed to the target variable which was associated with period *t*. In this way, we could show, in principle, how the history of the habitat can also be included as a predictor. Indeed, this is the first step at showing how the present methodology may be developed to include SIRs-type modelling properties.

## Data Availability

All data produced in the present study are available upon reasonable request to the authors. Epidemiological (confirmed cases of COVID-19) data is available at: https://www.gob.mx/salud/documentos/datos-abiertos-152127

## Acknowledgments

The EpI-PUMA project has benefited from the collaboration of multiple researchers, students and software developers. We are grateful for financial support from DGAPA-PAPIIT special project grant IV100520.

## Supplementary material

### Time dependent models

Here we offer more detail as to how time dependence is included into an SDM/ENM. We consider a time dependent variable of interest, *v*(*t*), defined by an epidemiological “who” ensemble, such as confirmed cases, deaths for age group *>* 60, mortality etc. The variable of interest may be ordinal, nominal or interval, and extensive or intensive. Its calculation requires a discretisation in time, so we may consider *N*_*T*_ time intervals, each of duration *δt*, to yield a total time period Δ_*T*_ = *N*_*T*_ *\* δt*. We may consider a variable *v*(*t, δt*) specified over an interval of time *δt*, such as the number of confirmed cases of COVID-19 in the *i*th time interval, *δt*_*i*_, or over the entire period, Δ_*T*_. We may further consider *v*_*α*_(*t, δt*), now calculated for a spatial cell *α* on a grid partition of *N*_*s*_ spatial cells on a region of interest. For instance, the number of confirmed cases in a country, *α*, in a month, *δt*, beginning at *t* being the first of June, 2021. Thus, (*α, δt*_*i*_) defines a spatio-temporal cell, to which we can assign a value of *v*(*t*), *v*(*α, δt*_*i*_). A target class *C* can be formed via any criterion that specifies a subset *S*_*C*_ *× 𝒯* _*C*_ *⊂ S × 𝒯* of cells. For instance, those spatial cells that have confirmed cases in the time interval *δt*_*i*_, or the 10% of spatial cells that have the highest mortality, as calculated in the interval *δt*_*i*_. Taking the discretisation (*α, δt*_*i*_) as the finest resolution, then *v*(Δ*S*, Δ*T*) can be calculated for any spatial region Δ*S* and time interval Δ*T*. In the case of an extensive variable, such as number of deaths, this corresponds to a simple aggregation.

Turning now to the habitat variables, **X**; in the same way, we can define a habitat variable *X*_*i*_(*α, δt*_*i*_) on a spatial cell in the time interval *δt*_*i*_. It may be that *X*_*i*_(*α, δt*_*i*_) = *X*_*i*_(*α*), i.e., it is independent of time over the interval of interest. For instance, over the lifetime of the pandemic we may take a climatic variable, such as average annual temperature, to be constant. On the other hand, if *X*_*i*_(*α, δt*_*i*_) represents the number of confirmed cases in the interval *δt*_*i*_, to be used as a niche variable, then certainly it will not be constant. The validity of an assumption of equilibrium depends on the rates of change and the magnitudes of change of *C* and **X** over the time period of interest. If the changes are slow and small over this period, then equilibrium may be a good approximation. Clearly, many variables of interest for COVID-19 are changing rapidly, such as vaccination rates, social distancing measures, travel restrictions etc.

Considering first a purely spatial model, we calculate *P* (*C*(*t*) | **X**(*t*)) using the formalism of section 4.2. This will be calculated using data for a period Δ_*T*_. A score for a niche variable 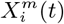 is given by 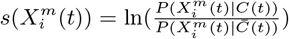, where 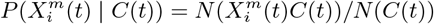 and 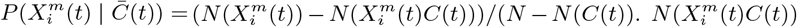 is the number of cells with a co-occurrence of 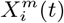 and *C*(*t*). For example, *C*(*t*) could represent the 10% of cells (Mexican municipalities) with the highest number of cases in month *t*, while 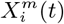 could represent a dynamic habitat variable, such as the 10% of municipalities with the highest average mobility in that month, or a more static variable, such as the 10% of municipalities with the highest average income, or those municipalities that have pixels from an average annual temperature raster in the range 20.6 *−* 22.3 deg *C*. 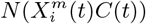 thus counts, for example, the number of municipalities that are in the top 10% with the highest number of cases in month *t* and that also are in the top 10% of municipalities with the highest intra-municipal daily labor flows. We can calculate 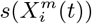 in different time intervals, *t*_*i*_, and compare 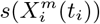 with 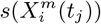. This can be done for all 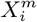 and the degree of correlation in time calculated.

Although 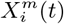 and *C*(*t*) may change in time, this does not imply that the ENM *P* (*C*(*t*) | **X**(*t*)) itself changes in time. The difference between 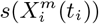 and 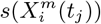, however, will inform us of any changes in the ENM itself. In the case that the niche is conserved, i.e., 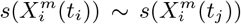, to predict the spatial distribution of the target class at *t*′, we use the scores, 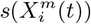, from the ENM at time *t*. For a given spatial cell, *α* at time *t*′ *> t*, we associate the score contribution 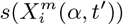. Thus, for example, if *X*_*i*_ is divided into deciles, 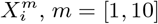, a given spatial cell at time *t* may be associated with decile *m*, whereas at time *t*′ it may be associated with a different decile *m*′, so that the cell *α* has become more niche-like/anti-niche-like in the cases that 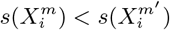 or *>* 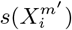 respectively.

Niche conservation implies that for a given habitat state, **X**, then *P* (*C*(*t*) | **X**) will not depend on time, i.e., there is the same probability to find the target species at any time *t*. When the niche is not conserved, however, then *P* (*C*(*t*) | **X**) ≠ *P* (*C*(*t*′) | **X**), i.e., the probability to find the target species given a certain habitat changes. In this case, the ENM *P* (*C*(*t*) | **X**(*t*)) will have limited predictability for *t*′ *> t*. To handle this situation we must only use *P* (*C*(*t*) | **X**(*t*)) to predict into the future with a time horizon Δ_*H*_ = (*t*′ *− t*) such that 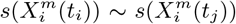. We may then create a new model at *t* + Δ_*H*_, *P* (*C*(*t* + Δ_*H*_) | **X**(*t* + Δ_*H*_)) and use it to predict at *t* + 2Δ_*H*_ and so on. In terms of a SDM, the fact that the habitat is changing means that spatial cells where the species was present/absent at *t* may now exhibit absence/presence. A useful way to quantify this, instead of using as target where the species is present in a given time period, is to focus on the *changes* between one period and the next. For example, we can take as target Δ_*C*_ (*t, t*′) those spatial cells where the species was present/absent in period *t* but then is absent/present in period *t*′. An explicit example is to consider those spatial cells where there were no confirmed cases in period *t −* 1 but there were cases in period *t*. The data for the pair of periods *t −* 1 and *t* is taken as training data and the corresponding scores calculated. The target class, 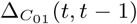, now consists of those spatial cells where there was an absence, *C*(*t −* 1) = 0, in period *t −* 1 and a presence, *C*(*t*) = 1, in period *t*.

Thus, for a habitat variable 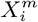 one calculates the score 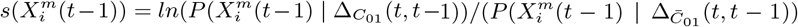, where 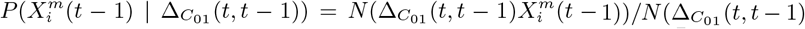 and 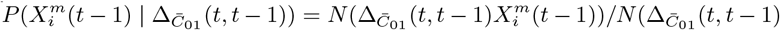, where 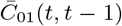 is the set complement of 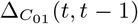. The scores 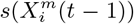 now define the ENM which can now be applied to the habitat variables 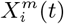 at time *t* to predict those cells that have an absence for confirmed cases at *t* but are predicted to have cases at *t* + 1. Similarly, for the class variable being top 10% of municipalities with highest number of cases, we may consider 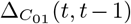 to represent those municipalities that were not in the top 10% at *t−*1 but, due to a worsening epidemiological situation, passed into the top 10% at time *t*.

Finally, although we will not enter into detail here, we may also consider as habitat variables, changes in the habitat. For example, we may consider 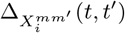, which represents those cells where there was a change in the habitat variable *X*_*i*_ from bin *m*′ to bin *m* passing from period *t*′ to period *t*.

## Footnotes

1 In the online system, EpI-PUMA, publicly available in a Platform-as-a-Service environment (http://covid19.c3.unam.mx), the above and many more different SDM/ENMs are available that use the methodology described in this paper.

2 In the EpI-PUMA system, publicly available in a Platform-as-a-Service environment (http://covid19.c3.unam.mx), 72 different SDM/ENMs are available that use the methodology described in this paper.

3 This is equivalent to the null hypothesis of type SIM2 in the classification of Gotelli [33]. This null hypothesis is one that leads to lower rates of Type I errors and corresponds, in the framework of presence-absence matrices, to keeping the number of observations fixed but randomising their location.

4 Although a complete factorisation of the likelihood may seem a strong assumption, the Naive Bayes approximation has been shown to be a robust performer even in cases where there are strong correlations between variables. An explanation for its surprisingly good performance can be found in [35].

